# Genetic and Proteomic Investigation of the Smoking-Parkinson’s Disease Association

**DOI:** 10.64898/2026.04.17.26351138

**Authors:** Mingjian Shi, Tommy Gunawan, Michael Setzer, Najeah Okashah, Yue Liu, Thomas S. Wingo, Aliza P. Wingo, Daniel Weintraub, Michael A. Schwarzschild, Christopher T. Rentsch, Henry R. Kranzler, Joshua C. Gray

## Abstract

**Background:** Epidemiological studies show an inverse association between cigarette smoking and Parkinson’s disease (PD), suggesting a potential protective effect of smoking on PD incidence, despite the well-established and overwhelming harms of smoking to human health. We integrated genomic and proteomic approaches to investigate the causality and molecular basis of this potential relationship.

**Methods:** We analyzed summary statistics from genome-wide association studies (GWAS) of smoking initiation (SmkInit), smoking intensity, and PD. Two-sample Mendelian randomization (MR) tested whether genetic liability to smoking behaviors causally influences PD risk. Shared genomic architecture was quantified using MiXeR, and conjunctional false discovery rate (conjFDR) analysis identified loci jointly associated with smoking and PD, which were then mapped to genes and tested for tissue enrichment. To identify mediating proteins, we integrated dorsolateral prefrontal cortex proteomic data with GWAS using proteome-wide association studies (PWAS), summary-based MR, heterogeneity in dependent instruments testing, and colocalization. Finally, the druggability of convergent genes was evaluated.

**Results:** MR analyses indicated a protective effect of genetic liability to SmkInit on PD risk (OR = 0.78, 95% CI: 0.67–0.91, *P* = 1.5 x 10^-3^), which was consistent across sensitivity analyses and not suggestive of directional pleiotropy. However, no significant effect of genetic liability to cigarettes per day (CigDay) on PD risk was found. MiXeR revealed modest polygenic overlap between SmkInit and PD (13.9%; genetic correlation *r_g_* = -0.16) and between CigDay and PD (22.9%; *r_g_* = -0.09). ConjFDR identified 95 shared loci for SmkInit-PD and 26 for CigDay-PD. SmkInit-PD loci mapped to genes involved in neurotrophic signaling, synaptic organization, microglial modulation, and mitochondrial stress responses, with expression enriched in substantia nigra, basal ganglia, and interconnected cortical regions. PWAS identified 11 proteins shared by PD and SmkInit and 5 shared with CigDay, several of which (AKT3, MAPT, RIT2, EXD2, and PPP3CC) were supported by both genomic and proteomic analyses. Druggability assessment highlighted six proteins with existing pharmacologic modulation potential, spanning neurotrophic, microglial, proteostatic, and ion-channel pathways.

**Conclusions:** Genetic liability to smoking initiation appears to confer modest protection against PD. Integrative genomic and proteomic evidence converges on neurotrophic, synaptic, microglial, and mitochondrial pathways as shared mechanisms, identifying biologically coherent potential therapeutic targets for advancing smoke-free neuroprotective strategies in PD.

## Introduction

Parkinson’s disease (PD) is a neurodegenerative disorder characterized by motor symptoms such as tremor, rigidity, bradykinesia, and postural instability, alongside a spectrum of non-motor symptoms such as cognitive impairment, psychiatric symptoms, sleep disturbances, and autonomic and sensory dysfunction [1, 2]. Epidemiologic studies consistently show an inverse association between cigarette smoking and PD risk, suggesting that smoking-related substituents confer neuroprotective benefits [3].

Smoking is associated with a reduced risk of PD relative to non-smoking, with stronger protective associations observed with greater smoking duration and intensity, and more pronounced among current than former smokers [4–7]. While biases such as selective mortality and underreporting have been examined [3, 8], these explanations do not fully account for the consistency of this inverse association across diverse populations and suggests that the possible biological mechanisms merit further inquiry.

Several mechanisms have been proposed to explain this epidemiological observation. PD primarily arises from the progressive loss of dopamine-producing neurons in the substantia nigra, a critical brain region involved in motor coordination and balance [9, 10]. Nicotine, the primary psychoactive compound in tobacco, activates nicotinic acetylcholine receptors (nAChRs), which regulate dopamine release, motor function, neuronal excitability, and cognitive functions [11]. Activation of these receptors has been suggested as one possible mechanism underlying the protective association between smoking and PD [12]. Experimental studies indicate that nicotine exposure enhances nAChR expression and activates intracellular signaling pathways that enhance neuronal resilience, particularly in brain regions susceptible to PD-related degeneration [13, 14]. However, a randomized controlled clinical trial using nicotine patches found no evidence that nicotine slows PD progression [15]. Beyond nicotine, smoking-induced inhibition of monoamine oxidase B (MAO-B), a mitochondrial enzyme involved in dopamine metabolism and generation of reactive oxygen species, has been proposed as a potential mechanism to explain smoking-associated PD protection [16, 17]. More recently, carbon monoxide (CO), another constituent of tobacco smoke, has been proposed to mediate smoking’s potentially neuroprotective effects [18, 19]. CO binds to hemoglobin with high affinity, reducing oxygen availability. At low doses that elevate carboxyhemoglobin to levels typical of smokers, CO has been shown in preclinical PD models to protect against oxidative stress and neurodegeneration [20, 21]. Consistent with these findings, a low-dose liquid formulation of CO is currently under clinical investigation for PD (clinicaltrials.gov identifier: NCT07005180). While biologically plausible, these candidate mechanisms have not been definitively established in humans.

Recent advances in statistical and molecular genetics provide a powerful framework to investigate the association and potential shared biology of smoking and PD. Genetic factors play a key role in PD susceptibility, with risk variants influencing mitochondrial function, protein homeostasis, and neuronal resilience to oxidative stress [22, 23]. Similarly, liability to tobacco smoking has a strong genetic component, with variants involved in nicotine metabolism, lung function, and nAChR and dopamine neuron function [24]. Leveraging these genetic architectures, methods such as Mendelian randomization (MR), polygenic overlap modeling, and conjunctional false discovery rate (conjFDR) enable rigorous tests of causality, directionality, and shared variant discovery between complex traits. Similar methodologies have clarified relationships among comorbid conditions such as body mass index (BMI) and problematic alcohol use [25], cannabis use and schizophrenia [26, 27], and depression and Alzheimer’s disease [28].

Complementary multi-omics approaches integrating genomic and proteomic data extend these approaches by connecting genetic risk to downstream molecular consequences, identifying cis-regulated brain proteins that mediate trait associations [29]. Integrated GWAS with brain proteogenomic data across smoking and PD allows for the detection of overlapping risk proteins and convergent molecular pathways, offering insight into the biological mechanisms that may connect smoking behavior and PD.

We applied this integrative framework to elucidate the molecular basis of the inverse relationship between smoking and PD. We utilized MR analyses to assess potential causal influences of genetic liability to smoking on PD risk. We assessed shared genomic architecture using MiXeR. Additionally, we applied conjFDR methods to identify shared genetic loci between smoking and PD, followed by gene mapping and tissue enrichment analyses to pinpoint relevant biological pathways. Finally, we conducted proteome-wide association studies (PWAS) of dorsolateral prefrontal cortex (DLPFC) tissue to identify cis-regulated brain proteins mediating shared genetic liability between PD and smoking, providing mechanistic context and identifying potential drug targets for PD.

## Methods

### Genome-wide Association Studies

Genome-wide association study (GWAS) summary statistics for smoking initiation (SmkInit) and cigarettes per day (CigDay) were obtained from the Sequencing Consortium of Alcohol and Nicotine Use (GSCAN) [24]. We used the SmkInit (N = 805,431) and CigDay (N = 326,497) datasets, consisting of individuals of European ancestry, after excluding 23andMe samples to comply with data-access restrictions. PD GWAS summary statistics were derived from the Global Parkinson’s Genetics Program (GP2) meta-analysis (GP2 & Leonard, 2025), excluding samples from the UK Biobank (UKB) and Million Veteran Program (MVP), comprising a total of 135,196 individuals (41,481 cases and 93,715 controls), combined with GWAS data from FinnGen Release 12 (R12) Parkinson’s Disease (G6_PARKINSON) dataset, which included 500,348 individuals (5,861 cases and 494,487 controls). These exclusions were implemented to minimize sample overlap between exposure and outcome GWAS and inflation by cross-trait enrichment. Thus, the final combined meta-analysis encompassed a total of 635,544 participants (Figure S1). Meta-analyses were conducted using METAL [31] with the inverse-variance weighting (IVW) method, which optimizes precision by weighting effect sizes according to their standard errors and adjusting for ancestry-specific sample sizes. The Uniformed Services University’s Human Research Protections Program Office classified this project as research not involving human subjects, in accordance with 32 CFR 219.102(e)(1) and applicable Department of War policy. All original studies had received ethical approval and obtained informed consent from all participants. A flowchart of the study design can be found in Figure 1.

**Figure 1.**
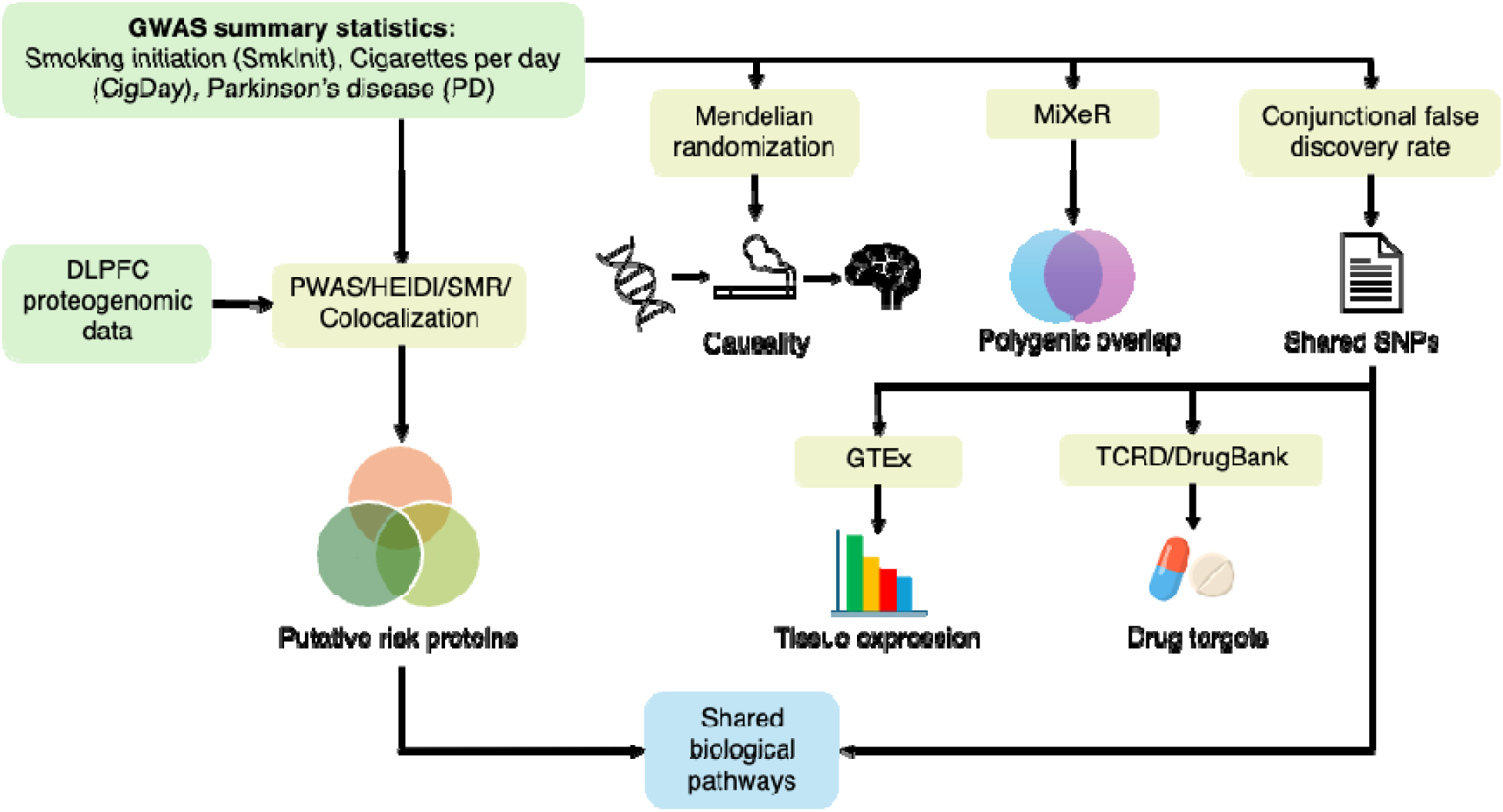
Study Workflow. Overview of the primary workflow performed in the study. Summary statistics from genome-wide association studies (GWAS) of SmkInit, CigDay, and PD were obtained and meta-analyzed. MR was used to test whether genetic liability for smoking behaviors causally influenced PD risk. Additional MR analyses using alternative smoking-related exposures (nicotine metabolite ratio) and positive control outcomes (lung cancer and coronary artery disease) were conducted but are not explicitly depicted for clarity. Polygenic overlap was quantified using MiXeR. Conjunctional false discovery rate (conjFDR) analysis identified loci jointly associated with smoking and PD, which were then used for Genotype-Tissue Expression (GTEx) analysis. Druggability of convergent genes was evaluated by referencing the Target Central Resource Database (TCRD) and DrugBank. To identify mediating proteins, we integrated dorsolateral prefrontal cortex (DLPFC) proteomic data with GWAS using protein-wide association study (PWAS), heterogeneity in dependent instruments (HEIDI), summary-based Mendelian randomization (SMR), and colocalization.

Other GWAS summary statistics of smoking-related exposures were obtained from publicly available GWAS of nicotine metabolite ratio (NMR) in current smokers (N = 5,185) [32] from the GWAS Catalog (https://www.ebi.ac.uk/gwas/), lung cancer (N = 40,187) [33], and coronary artery disease (CAD; N = 628,001) from the CARDIoGRAMplusC4D Consortium (https://cardiogramplusc4d.org/) [34]. NMR, defined as the ratio of cotinine to 3-hydroxycotinine, serves as a biomarker of nicotine metabolism rate and reflects CYP2A6 enzymatic activity [35]. Higher NMR values indicate faster nicotine metabolism and lower systemic nicotine exposure per cigarette. Since NMR is only assessed in current smokers, we included it as a test of whether differences in nicotine metabolism, and by extension nicotine exposure, causally influence PD risk, separate from SmkInit and CigDay.

### Mendelian Randomization Analysis

We conducted two-sample MR analyses to assess potential causal relationships between smoking-related traits and PD using large GWAS summary statistics (Hartwig et al., 2016). Independent instrumental variables were selected using genome-wide significant SNPs (*P* < 5 × 10) for each trait, followed by linkage disequilibrium clumping (*r*² < 0.01, window size 10,000 kb) to ensure independence. Palindromic and ambiguous SNPs were excluded to prevent strand mismatches, and harmonization procedures aligned effect alleles across exposure and outcome datasets using the 1000 Genomes Project reference panel.

We evaluated three smoking-related phenotypes: SmkInit, CigDay representing smoking intensity, and NMR, a biomarker of nicotine metabolism. To validate the reliability of our MR design, we incorporated positive control outcomes including lung cancer and CAD, which have established causal relationships with smoking [37]. MR analyses assessed the effects of each exposure on PD risk, with additional analyses examining smoking exposures on lung cancer or CAD to confirm expected positive associations.

Primary causal estimates were derived using the inverse-variance weighted (IVW) method, which assumes no directional pleiotropy. To assess robustness and account for potential pleiotropy violations, we implemented complementary methods including MR-Egger regression [38], weighted median, and MR-PRESSO (Mendelian Randomization Pleiotropy RESidual Sum and Outlier) [39] to identify and remove pleiotropic outlier SNPs. Additionally, we applied Mendelian Randomization Accounting for Pleiotropy and Sample Structure (MR-APSS), which improves causal inference by adaptively selecting valid instruments while accounting for polygenicity and sample structure [40]. All MR analyses were conducted in R using the TwoSampleMR [41], MRPresso [39], and MR-APSS packages [40].

### Characterizing Polygenic Overlap

To quantify the shared genetic architecture between SmkInit and PD as well as CigDay and PD, we employed MiXeR [42], a polygenic mixture model. We first fitted univariate MiXeR models for each trait to estimate its polygenicity, defined as the number of SNPs accounting for 90% of SNP-based heritability, and its discoverability, which represents the average effect size of causal variants. The statistical power and suitability of each univariate model were confirmed using the Akaike Information Criterion (AIC). Afterwards, bivariate MiXeR analyses were conducted to estimate the number of unique and overlapping causal variants between SmkInit and PD. These models also determined the proportion of shared variants exhibiting concordant directions of effect. The overall extent of polygenic overlap was quantified using the Dice coefficient implemented in MiXeR. To visualize cross-trait enrichment, we generated conditional quantile-quantile (Q-Q) plots, which show the distribution of association *P*-values for one phenotype across significance strata (*P* ≤ 0.1, 0.01, and 0.001) of the other.

### Conjunctional False Discovery Rate (conjFDR) Analysis

We conducted conjunctional false discovery rate (conjFDR) analysis [43] to identify loci jointly associated with SmkInit and PD as well as CigDay with PD. Conditional FDR (condFDR) values were calculated bidirectionally, first conditioning PD on the smoking phenotype and then conditioning the smoking phenotype on PD. The larger of the two condFDR estimates was set as the conjFDR value, ensuring a conservative threshold for joint associations. Loci with conjFDR < 0.05 were considered significantly shared.

### Genomic loci definition and gene-set enrichment

ConjFDR significant SNPs were clumped with PLINK v1.90b7.7 (64-bit) to identify LD-independent lead SNPs/loci. Independent significant SNPs were identified using the European ancestry 1000 Genomes reference and an LD lock distance criteria for merging (≤250 kb, *r*^2^ < 0.6). For each locus, only lead SNPs (*r*^2^ < 0.1) with the smallest conjFDR values were chosen. Lead SNPs that were not genome-wide significant (*P* < 5 × 10^-8^) in the SmkInit, CigDay, and PD summary statistics were considered novel. Lead SNPs were mapped to genes based on their genomic location, specifically by overlap within gene boundaries or proximity to the nearest gene. BiomaRt (Durinck et al., 2009) and ANNOVAR (Wang et al., 2010) was used to map the nearest gene. When the two disagreed, we verified coordinates in dbSNP (https://www.ncbi.nlm.nih.gov/snp/). Gene expression and tissue specificity analyses were performed in FUMA using Genotype-Tissue Expression (GTEx), with expression levels normalized as transcripts per million to account for differences in library size and sequencing depth. Multiple testing corrections for the enrichment analyses were applied using FDR adjustment.

### Brain Proteomic Data

The human genetic and DLPFC proteomic data used for PWAS and Mendelian randomization (SMR) [44] were collected through collaborations involving the AMP-AD [45] and AMP-AD Diversity [46] initiatives across multiple research sites. All donors or their next of kin provided informed consent and repository approval, allowing their data and biospecimens to be utilized for research. Genetic information for donor samples was obtained from brain tissue or blood through whole-genome sequencing (WGS) or array-based phenotyping, as previously described [29]. Brain samples were collected for proteomic analysis and profiled using tandem mass tag (TMT) mass spectrometry, as previously reported [29, 46, 47]. The DLPFC was selected as it was a region available in our postmortem cohorts. DLPFC protein expression has been shown to correlate with other cortical regions, supporting its use as a representative cortical tissue for large-scale proteomic analyses [48, 49]. Quality control was conducted as previously described in detail (Wingo et al., 2023; Wingo et al., 2022), resulting in a normalized proteomic profile of 9,725 proteins from which biological (sex, age at death, clinical diagnosis), technical (batch, PMI), and hidden confounding variables have been removed.

### Identification of putative risk proteins

We accessed the GWAS summary statistics for smoking phenotypes [24] and PD (GP2 & Leonard, 2025) derived from the same non-overlapping cohorts used in our primary MR analyses. PWAS was conducted using the FUSION pipeline, integrating GWAS summary statistics with brain proteomic data [52]. The PWAS identified risk proteins whose genetically-controlled abundance level is linked to smoking behaviors and PD, respectively, with significance defined as FDR < 0.05. To determine whether these risk proteins mediate the SNP-trait relationship, we performed summary-based Mendelian randomization (SMR) [44] on the risk proteins identified in the PWAS. Brain protein mediation of the association between the SNP and trait can arise from causality, pleiotropy, or linkage disequilibrium. Heterogeneity in dependent instruments (HEIDI) testing was used to check whether associations were due to linkage disequilibrium [44]. Proteins with HEIDI *P* < 0.05 were assumed to have an association due to linkage disequilibrium and excluded from causal candidates. For the colocalization test, we used the COLOC software [53] to estimate the posterior probability of the protein and phenotypes sharing a causal variant (PP4), and the posterior probability of the protein and phenotypes not sharing a causal variant using marginal association statistics. The final candidate risk proteins had a PWAS FDR < 0.05, an SMR *P*-value < 0.05, and a HEIDI *P*-value > 0.05, or had a PWAS FDR < 0.05 and PP4 > 0.75 in COLOC.

### Drug Repurposing

We queried loci identified from conjFDR through the Target Central Resource Database (TCRD; https://pharos.nih.gov/) to identify potential therapeutic targets, druggability, and drug-protein interactions. TCRD classifies targets based on druggability: approved drugs with known mechanisms (Tclin); drug activity meeting specific thresholds (Tchem); biological targets with no known drugs but with Gene Ontology (GO) leaf term annotations or Online Mendelian Inheritance in Man (OMIM) phenotypes, or meeting at least two of the following three conditions: a fractional PubMed count >5, >3 National Center for Biotechnology Gene Reference Intro Function annotations, or >50 commercial antibodies (Tbio); and proteins curated in UniProt, but lacking substantial research or druggability criteria (Tdark) [54]. For targets classified as Tclin in TCRD, we queried DrugBank version 5.1.13 (https://go.drugbank.com) to characterize the identified drugs and their therapeutic indications.

## Results

### MR Results

We observed a significant protective association between genetic liability for SmkInit and PD (Figure 2). The IVW method detected a significant protective effect (OR = 0.78, 95% CI: 0.67–0.91, *P* = 1.5 × 10⁻³), suggesting that genetic susceptibility to SmkInit is associated with decreased PD risk. This finding was supported by the weighted median method (OR = 0.73, 95% CI: 0.62–0.87, *P* = 5.0 × 10⁻□), MR-APSS (OR = 0.94, 95% CI: 0.89–0.99, *P* = 0.027) and MR-Egger regression (OR = 0.47, 95% CI: 0.25–0.90, *P* = 2.3 × 10⁻²). The MR-Egger intercept was not significant (intercept = 0.0063, *P* = 0.114), consistent with no directional pleiotropy. Although MR-PRESSO detected horizontal pleiotropy (RSS_obs_ = 628.02, *P* < 6.67 × 10⁻□), the distortion test showed that outlier removal had minimal impact on the causal estimate (distortion *P* = 0.65) (Table S1).

**Figure 2.**
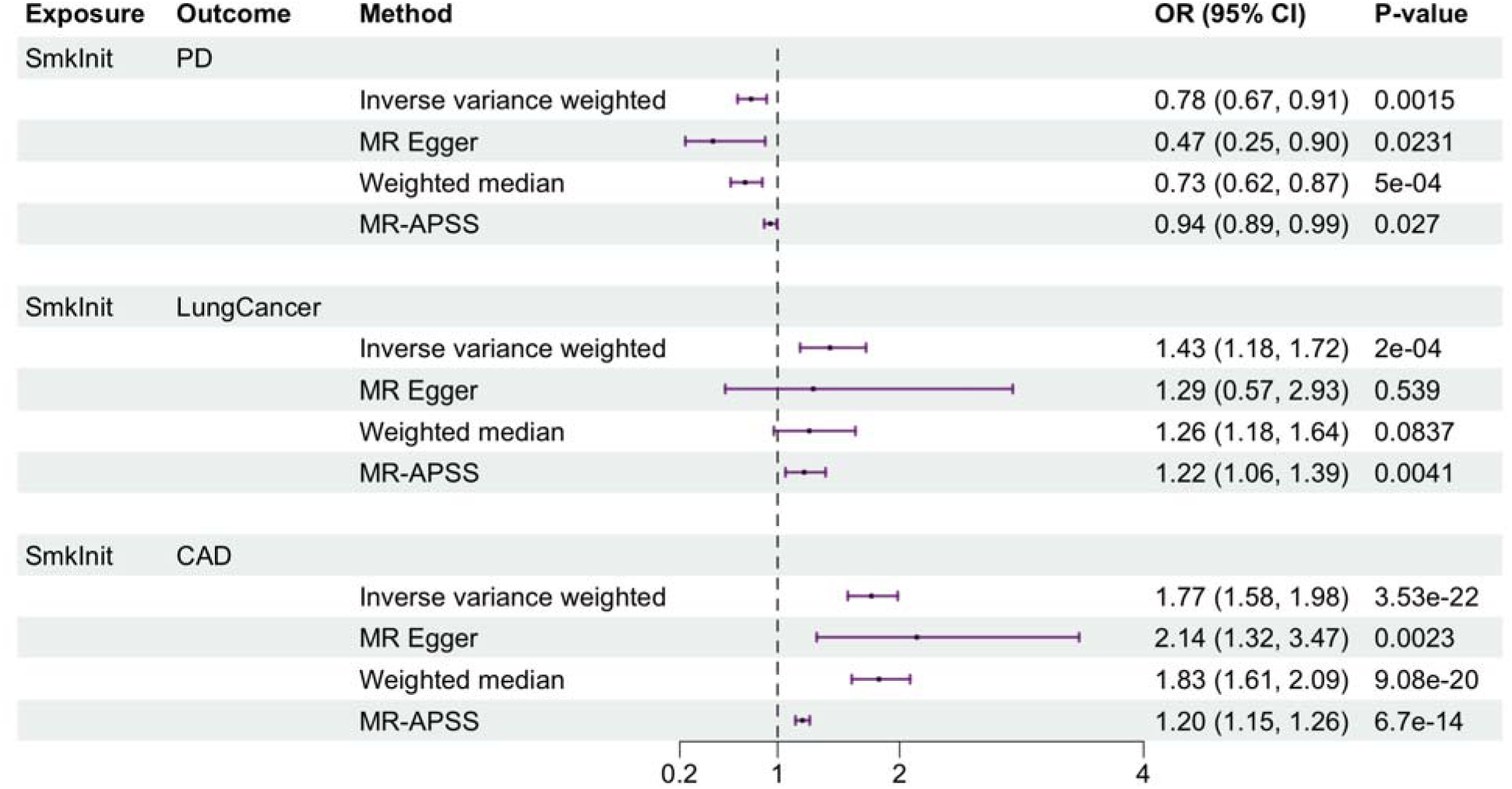
Mendelian randomization (MR) of SmkInit and disease risk. Forest plots show odds ratios (OR) with 95% confidence intervals from four MR estimators, Inverse Variance Weighted (IVW), MR-Egger, Weighted Median, and MR-APSS. The dashed vertical line marks the null (OR = 1). Panels depict the effect of SmkInit as exposure on three outcomes: PD (top), lung cancer (middle), and coronary artery disease (CAD) (bottom). Points indicate OR estimates; horizontal bars show 95% CIs.

As positive controls, we confirmed well-established causal relationships between smoking and the outcomes of CAD and lung cancer (Figure 2). IVW analysis revealed a strong positive association of SmkInit with CAD, (OR = 1.77, 95% CI: 1.58–1.98, *P* = 3.53 × 10⁻²²), consistently supported across all sensitivity analyses. IVW analysis also demonstrated that genetic liability to SmkInit significantly increased lung cancer risk (OR = 1.43, 95% CI: 1.18–1.72, *P* = 2.0 × 10⁻□), with additional support from MR-APSS (OR = 1.22, 95% CI: 1.06–1.39, *P* = 4.1 × 10⁻³). While weighted median and MR-Egger analyses showed a consistent direction of effect, they did not reach statistical significance (OR = 1.26, 95% CI: 0.97–1.64, *P* = 0.084; and OR = 1.29, 95% CI: 0.57–2.93, *P* = 0.539, respectively).

We also investigated whether the genetic liability for smoking intensity conferred a protective effect against PD. Using CigDay as exposure and PD as outcome, IVW analysis revealed a non-significant association (OR = 0.90, 95% CI: 0.75–1.08, *P* = 0.254) that was consistent across other methods: MR-Egger (OR = 0.94, 95% CI: 0.68–1.28, *P* = 0.687), weighted median (OR = 0.98, 95% CI: 0.81–1.19, *P* = 0.842), and MR-APSS (OR = 0.98, 95% CI: 0.89–1.07, *P* = 0.628). As a positive control, CigDay showed a robust association with lung cancer in IVW analysis (OR = 8.83, 95% CI: 6.33–12.31, *P* = 8.99 × 10⁻³), which was supported across all MR methods (Figure S2). However, no significant relationship was found between CigDay and CAD using IVW (OR = 1.20, 95% CI: 0.96–1.49, *P* = 0.11) or any of the other MR methods (MR-Egger: OR = 0.83, 95% CI: 0.59–1.17, *P* = 0.283, weighted median: OR = 0.99, 95% CI: 0.86–1.14, *P* = 0.917, MR-APSS: OR = 1.01, 95% CI: 0.92–1.13, *P* = 0.757). Finally, NMR showed no evidence of a causal relationship with PD across any method: IVW (OR = 1.00, 95% CI: 0.98–1.03, *P* = 0.748), MR-Egger (OR = 1.00, 95% CI: 0.95–1.06, *P* = 0.95), weighted median (OR = 1.01, 95% CI: 0.99–1.03, *P* = 0.365), and MR-APSS (OR = 1.00, 95% CI: 0.99–1.01, *P* = 0.693) (Figure S3). Collectively, these MR analyses support a causal relationship of genetic liability to SmkInit and PD risk, but not CigDay and PD risk. Due to the greater statistical power of the SmkInit GWAS, we focus on the SmkInit results here and provide detailed CigDay results in the Supplementary Results.

### Shared Genomic Architecture (MiXeR)

Univariate MiXeR models confirmed adequate power for subsequent bivariate analyses, as evidenced by finite AIC values, reflecting good model fit. MiXeR analysis revealed a mild negative genetic correlation (*r_g_*= -0.156) between SmkInit and PD, with a polygenic overlap quantified by a Dice coefficient of 13.9%. Specifically, of an estimated 4,329 potential causal variants linked to SmkInit and 994 associated with PD, approximately 369 variants were shared. Notably, only 14.2% of the shared causal variants displayed concordant effect directions (Table S2).

Conditional Q-Q plots demonstrated cross-trait genetic enrichment between SmkInit and PD, further supporting shared polygenic architecture. In these analyses, SNPs were stratified by their association with the primary trait (e.g., PD) at increasingly stringent p-value thresholds (*P* ≤ 0.1, 0.01, and 0.001). As shown in the conditional Q-Q plots, SNPs more strongly associated with PD exhibited progressively greater enrichment for association with SmkInit, illustrated by an increasingly leftward shift from the null line. The same reciprocal enrichment pattern was observed when stratifying by SmkInit significance and evaluating PD associations. These non-random enrichment patterns confirm that a subset of shared genetic variants influence both traits, reinforcing the polygenic overlap detected by MiXeR (Figure 3).

**Figure 3.**
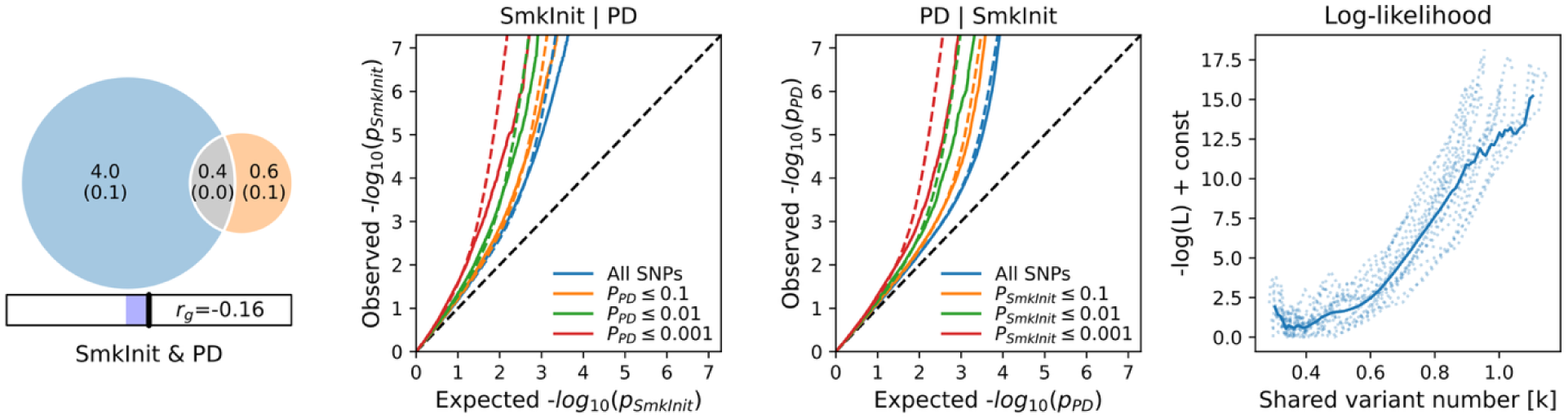
SmkInit-PD bivariate MiXeR results. The Venn diagram (left) illustrates the estimated numbers of causal genetic variants (in thousands) unique to SmkInit (blue), unique to PD (orange), and variants shared between both traits (gray). Genetic correlation (*r_g_*) is indicated beneath. The conditional Q–Q plots (middle) demonstrate cross-trait enrichment, showing increased polygenic signals for SmkInit conditional on PD (SmkInit|PD) and for PD conditional on SmkInit (PD|SmkInit), across varying thresholds of SNP significance (*P* ≤ 0.1, 0.01, 0.001). The right panel shows the log-likelihood curve used to estimate the number of shared causal variants, illustrating model fit across variant number estimations.

### Shared genetic loci (cond/conjFDR)

At conjFDR < 0.05, we identified 95 significant loci associated with both SmkInit and PD. Of these, 48 variants were not genome-wide significant in the individual SmkInit summary statistics, and 57 were not significant in the PD summary statistics. Of the lead SNPs, 52 (59%) had concordant (consistent) effect directions for SmkInit and PD, and 43 had discordant (opposite) effect directions (Table S3 & Figure S4). The implicated genes converge on microglial and neuronal biology, including the microglial purinergic receptor gene *P2RY12*; PI3K-AKT neurotrophic signaling pathway (*NTRK2*/*BDNF*, *AKT3*); synaptic vesicle/scaffolding components (*BSN*, *RIT2*, *NTM*); mitochondrial/oxidative and DNA-repair stress signaling (*EXD2*, *ERCC8*, *MSRA*, *MCRS1*); and established PD-relevant neurodegeneration genes (*MAPT*, *DYRK1A*, *APP*) (Table S3).

### FUMA Genotype-Tissue Expression analysis

Analysis of GTEx data for genes linked to the 95 lead SNPs associated with both SmkInit and PD showed significant tissue-specific expression enrichment after Bonferroni correction. In the 54-tissue panel, signals were concentrated in PD-relevant brain regions, including the basal ganglia (putamen, caudate, nucleus accumbens), hypothalamus, hippocampus, amygdala, anterior cingulate cortex (BA24), frontal cortex (BA9), substantia nigra, and cerebellum, with additional enrichment in tibial nerve, liver, heart (left ventricle), pancreas, uterus, stomach, and adrenal gland (Figure 4). At the level of 30 general tissues, enrichment was observed for nervous tissue and several peripheral tissues, most notably heart, liver, kidney, pancreas, uterus, muscle, stomach, adrenal gland, and blood/vessel (Figure S5).

**Figure 4.**
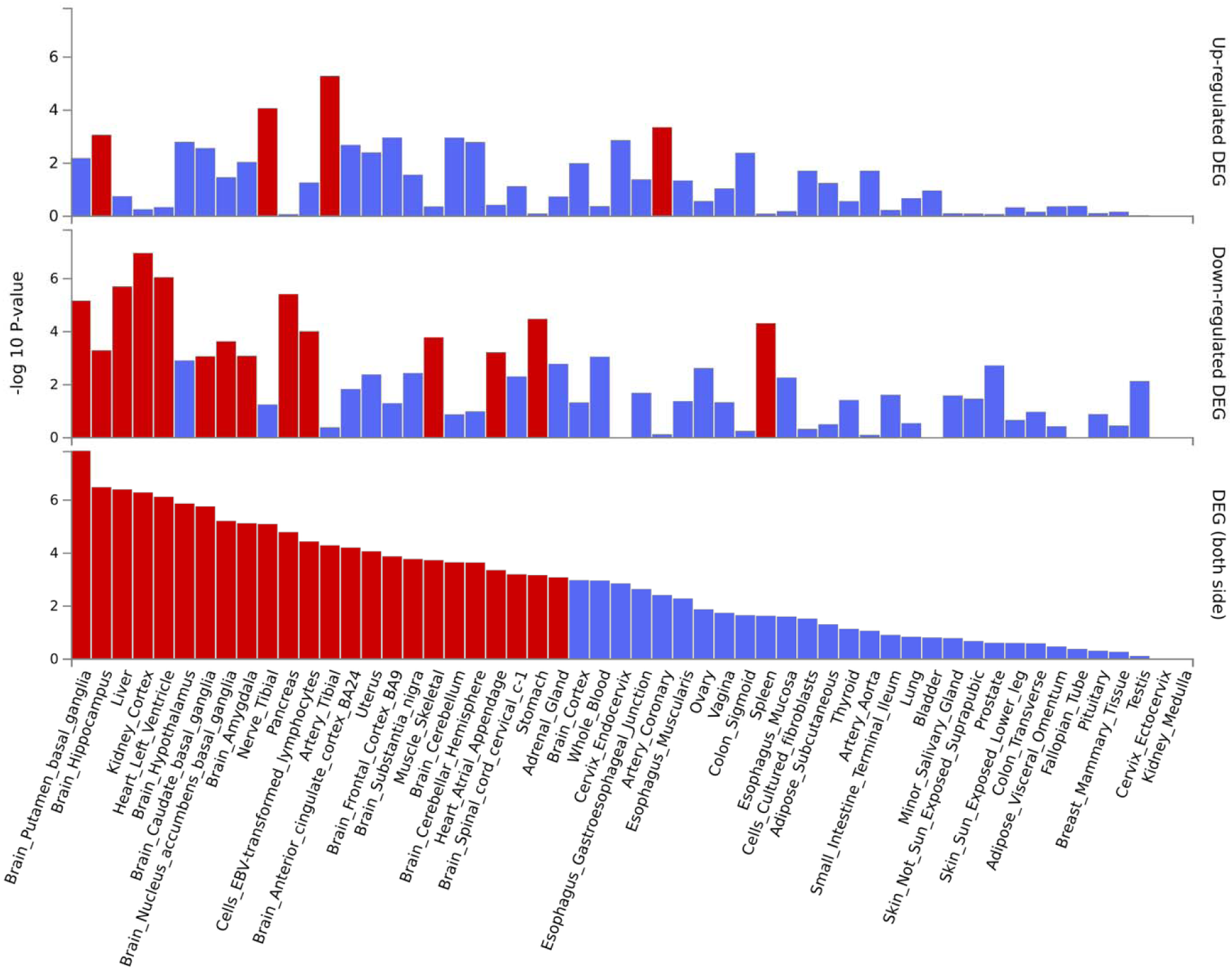
Differential gene expression (DEG) in 54 GTEx tissue types for genes linked to lead SNPs in distinct loci significantly associated with SmkInit and PD. Significant enrichment (*P* < 0.05 after Bonferroni correction, two-sided tests) is highlighted in red.

### Integrating PD GWAS with brain proteomes

PWAS followed by SMR and HEIDI identified 214 putative risk proteins for SmkInit, 76 for CigDay, and 117 for PD (Table S5-S7). Cross-trait comparison revealed 11 overlapping risk proteins between SmkInit and PD (Table 1), and 5 overlapping causal proteins between CigDay and PD (Table S8). Notably, 7 of the 11 overlapping proteins between SmkInit and PD showed discordant directions of association (Table 1), such that genetically increased protein abundance was associated with increased risk for one trait but decreased risk for the other. This pattern suggests that these proteins reflect the biology underlying the inverse relationship between SmkInit and PD.

**Table 1.**
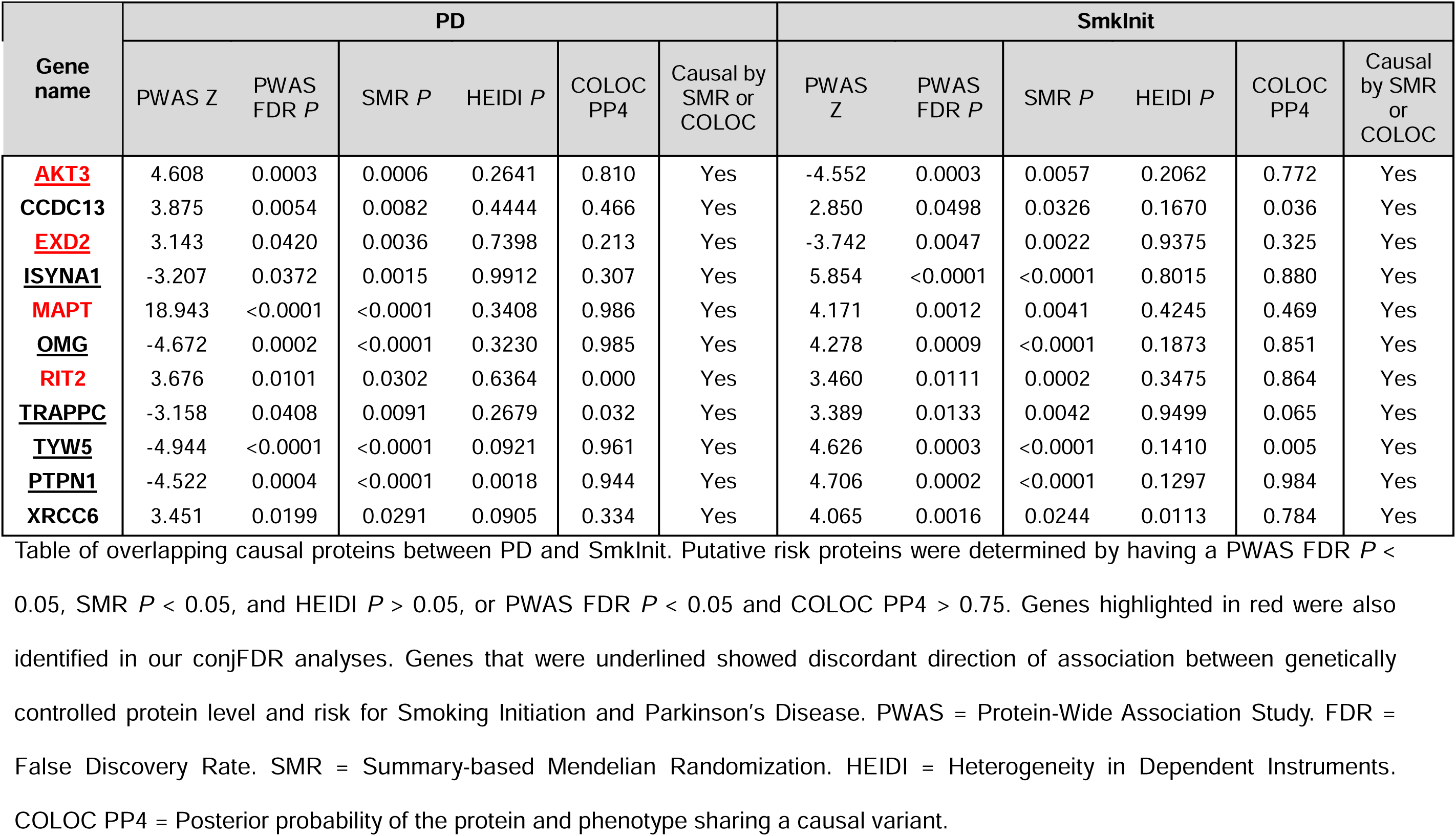
Overlapping putative risk proteins from PWAS of PD and SmkInit.

| Gene name | PD |  |  |  |  |  | Smklnit |  |  |  |  |  |
| --- | --- | --- | --- | --- | --- | --- | --- | --- | --- | --- | --- | --- |
|  | PWAS Z | PWAS FDR *P* | SMR *P* | HEIDI *P* | COLOC PP4 | Causal by SMR or COLOC | PWAS Z | PWAS FDR *P* | SMR *P* | HEIDI *P* | COLOC PP4 | Causal by SMR or COLOC |
| **AKT3** | 4.608 | 0.0003 | 0.0006 | 0.2641 | 0.810 | Yes | -4.552 | 0.0003 | 0.0057 | 0.2062 | 0.772 | Yes |
| **CCDC13** | 3.875 | 0.0054 | 0.0082 | 0.4444 | 0.466 | Yes | 2.850 | 0.0498 | 0.0326 | 0.1670 | 0.036 | Yes |
| **EXD2** | 3.143 | 0.0420 | 0.0036 | 0.7398 | 0.213 | Yes | -3.742 | 0.0047 | 0.0022 | 0.9375 | 0.325 | Yes |
| **ISYNA1** | -3.207 | 0.0372 | 0.0015 | 0.9912 | 0.307 | Yes | 5.854 | <0.0001 | <0.0001 | 0.8015 | 0.880 | Yes |
| **MAPT** | 18.943 | <0.0001 | <0.0001 | 0.3408 | 0.986 | Yes | 4.171 | 0.0012 | 0.0041 | 0.4245 | 0.469 | Yes |
| **OMG** | -4.672 | 0.0002 | <0.0001 | 0.3230 | 0.985 | Yes | 4.278 | 0.0009 | <0.0001 | 0.1873 | 0.851 | Yes |
| **RIT2** | 3.676 | 0.0101 | 0.0302 | 0.6364 | 0.000 | Yes | 3.460 | 0.0111 | 0.0002 | 0.3475 | 0.864 | Yes |
| **TRAPPC** | -3.158 | 0.0408 | 0.0091 | 0.2679 | 0.032 | Yes | 3.389 | 0.0133 | 0.0042 | 0.9499 | 0.065 | Yes |
| **TYW5** | -4.944 | <0.0001 | <0.0001 | 0.0921 | 0.961 | Yes | 4.626 | 0.0003 | <0.0001 | 0.1410 | 0.005 | Yes |
| **PTPN1** | -4.522 | 0.0004 | <0.0001 | 0.0018 | 0.944 | Yes | 4.706 | 0.0002 | <0.0001 | 0.1297 | 0.984 | Yes |
| **XRCC6** | 3.451 | 0.0199 | 0.0291 | 0.0905 | 0.334 | Yes | 4.065 | 0.0016 | 0.0244 | 0.0113 | 0.784 | Yes |

Table of overlapping causal proteins between PD and Smklnit. Putative risk proteins were determined by having a PWAS FDR *P* < 0.05, SMR *P* < 0.05, and HEIDI *P* > 0.05, or PWAS FDR *P* < 0.05 and COLOC PP4 > 0.75. Genes highlighted in red were also identified in our conjFDR analyses. Genes that were underlined showed discordant direction of association between genetically controlled protein level and risk for Smoking Initiation and Parkinson's Disease. PWAS = Protein-Wide Association Study. FDR = False Discovery Rate. SMR = Summary-based Mendelian Randomization. HEIDI = Heterogeneity in Dependent Instruments. COLOC PP4 = Posterior probability of the protein and phenotype sharing a causal variant.

Among these discordant proteins, AKT3 and EXD2, which were also identified in the conjFDR analyses, map onto neurotrophic and mitochondrial stress-response pathways, respectively. AKT3 is a core component of the PI3K-AKT-mTOR signaling pathway implicated in neuronal survival and plasticity [55], and EXD2 regulates mitochondrial homeostasis and oxidative stress response pathway (Silva et al., 2018). The additional discordant proteins (ISYNA1, OMG, TYW5, PTPN1, XRCC6) implicate metabolic, myelination, and DNA repair pathways [57–61], underscoring the heterogeneity of antagonistic pathways underlying SmkInit and PD.

In contrast, a smaller subset of overlapping proteins, including MAPT and RIT2, showed concordant effects between SmkInit and PD. MAPT is involved in cytoskeletal integrity and tau regulation [62], and RIT2 in synaptic vesicle trafficking and dopaminergic neurotransmission. Together, these results highlight that the relationship between smoking and PD is driven by a mixture of discordant and concordant protein-level mechanisms, with discordant effects representing key molecular signature of antagonistic pleiotropy.

### Drug repurposing

We identified six genes from the conjFDR analyses that were classified as Tclin (*APP*, *KCNH5*, *MAPT*, *NTRK2*, *P2RY12*, and *XPO1*), indicating that they interact with FDA-approved medications and are potential targets for drug repurposing (Table S9). *APP* encodes the amyloid-beta precursor protein, whose metabolite amyloid-beta is a hallmark of Alzheimer’s disease. While monoclonal antibody aducanumab was identified as a potential medication for repurposing, it has been withdrawn from the market and has been replaced by newer generation amyloid immunotherapy lecanemab and donanemab [63, 64]. Retinol and its derivative tretinoin were also identified as candidates for repurposing and may reduce oxidative stress and neuroinflammation associated with PD [65]. *KCNH5*, encoding the voltage-gated potassium channel protein Kv10.2, is linked with epilepsy and neurodevelopmental disorders [66] and is modulated by potassium channel blockers fampridine, amifampridine, guanidine, and quinidine. *MAPT* is a well-established risk gene for PD and is essential for neuronal structure and transport [67] and interacts with the proton-pump inhibitor lansoprazole. *NTRK2* encodes tropomyosin receptor kinase B (TrkB), a key receptor for brain-derived neurotrophic factor that regulates neuronal development, function, and survival [68]. Several clinically approved NTRK/ALK inhibitors (crizotinib, bosutinib, nintedanib, lortlatinib, larotrectinib, entrectinib) represent opportunities for testing TrkB-mediated neurotrophic modulation in PD. Lastly, *XPO1* encodes exportin 1, which transports proteins and RNAs from the nucleus to the cytoplasm [69], which together with the nuclear export inhibitor selinexor suggests that XPO1-mediated mechanisms are relevant to PD and warrant further investigation.

## Discussion

Despite consistent epidemiological evidence of an inverse association between cigarette smoking and PD, the mechanisms of this effect remain a puzzle [3]. Through convergent evidence across MR, polygenic architecture modeling, cross-phenotype locus discovery, and proteomic analyses, our study provides a clearer picture of the potential causal and biological basis of this relationship. Rather than implicating a single pathway, our results map the smoking–PD relationship onto an array of genes and proteins implicated in neuronal survival, synaptic organization, and microglial modulation [70, 71].

Our MR analyses provide evidence that genetic liability to SmkInit confers modest protection against PD. This protective association was consistent across multiple MR estimators, without evidence of directional pleiotropy, and contrasts with the expected positive associations with CAD and lung cancer (Larsson et al., 2020). These patterns suggest, but do not definitively establish, that the observed protective effect on PD risk may include a causal component beyond residual confounding or pleiotropic influences. Any potential protective association with PD is far outweighed by the substantial harms of smoking [37].

In contrast to SmkInit, genetic liability for CigDay and NMR showed no causal associations with PD. Although CigDay robustly predicted lung cancer, its association with CAD was null, consistent with the idea that CigDay reflects peak smoking intensity among current smokers rather than cumulative lifetime exposure, which more strongly influences vascular outcomes [73]. Notably, genetic liability to NMR, which indexes nicotine clearance and systemic nicotine exposure, showed no evidence of a causal association with PD risk. These findings align with randomized clinical trial evidence indicating that nicotine administration does not slow PD progression [15]. Importantly, this randomized clinical trial evaluated the role of nicotine in disease progression rather than disease onset and does not exclude the possibility that nicotine could influence early neurodegenerative processes in populations at-risk for PD. However, the null MR findings for MR reduces support for nicotine as a plausible preventative agent.

These findings diverge from epidemiological findings that greater smoking intensity confers stronger protection against PD [3, 6, 74]. Rather than a dose-dependent effect of tobacco smoke exposure on PD risk, our results suggest that these observational patterns may reflect bias. While SmkInit, is influenced by social and environmental factors such as cigarette availability [75], it captures an upstream liability to initiate smoking. In contrast, CigDay and NMR are conditional phenotypes measured only among individuals who have smoked. Thus, they are susceptible to index-event and selection biases, including differential cessation, survival, and nonrandom participation. These biases can generate spurious or attenuated associations with PD, helping to explain why observational studies sometimes report inverse CigDay-PD relationships, whereas our MR analyses indicate no causal effect of CigDay on PD despite strong positive effects on lung cancer.

Our integrated genetic analyses shed light on plausible mechanisms underlying the relationship between SmkInit and PD. We found a modest inverse genetic correlation between SmkInit and PD (*r_g_* ≈ -0.16) and a limited polygenic overlap (Dice coefficient ≈ 14%). More strikingly, among this shared subset, the majority of variants (≈86%) were discordant in their direction of effects, indicating that most alleles that increase the likelihood of smoking initiation decrease PD risk and vice versa. Thus, rather than a broad, genome-wide protective effect, the relationship between the two traits appears to be driven by a small, mechanistically focused genetic components with mixed directional effects.

Using conjFDR analysis, we identified 95 shared loci between SmkInit and PD. Effect directions were heterogeneous, with 52 loci (59%) showing concordant effects and 43 (41%) showing discordant effects, consistent with antagonistic pleiotropy. These loci converged on several biologically coherent pathways related to microglial function, neuronal survival, synaptic maintenance, and mitochondrial response, key processes in PD pathogenesis. Microglial signaling was represented by *P2RY12*, a purinergic receptor expressed on microglia that regulates activation states and neuroinflammatory responses [76, 77]. This finding is consistent with the role of microglial activation in PD pathogenesis [78, 79]. Neurotrophic and survival signaling pathways were implicated by *NTRK2* (encoding the TrkB receptor for BDNF) and *AKT3*, core components of the PI3K-AKT cascade that promotes neuronal survival, growth, and stress resistance that has been implicated in PD [55, 80, 81]. Synaptic scaffolding and neurotransmission were represented by *BSN*, which maintains presynaptic terminal integrity and neurotransmission that are disrupted in PD [82]. Mitochondrial stress responses, involving *EXD2*, protects the mitochondria from oxidative injury. Finally, canonical PD risk genes *MAPT*, *DYRK1A*, and *APP* highlight the roles of cytoskeletal dynamics and protein aggregation in PD [83]. Additionally, several of the loci identified (*CNNM2*, *KANSL1*, *MAPT*, *NSF*, *SPPL2C*) were also reported in a recent large cross-disorder study of complex neurological and psychiatric traits [84]. Their recurrence across independent datasets reinforces the biological relevance of the loci identified and supports our interpretation that the relationship between smoking and PD is driven by pathway-specific pleiotropy. Several genes identified from our conjFDR analyses (*CTSB*, *DYRK1A*, *RIT2*, *SCARB2*, and *TMEM163*) were also prioritized as druggable targets by their polygenic priority scores [85]. Together, these findings outline a network of microglial, neurotrophic, synaptic, and mitochondrial pathways that shape PD risk and present opportunities for PD therapeutic development.

To determine whether these genetic effects extend to the protein level, we conducted PWAS followed by SMR and HEIDI integrating GWAS and DLPFC proteomic data, which identified 11 overlapping risk proteins between SmkInit and PD. Notably, four of the proteins, AKT3, MAPT, RIT2, and EXD2, were also identified from our conjFDR analyses. Two of these proteins, AKT3 and EXD2, showed discordant effects between SmkInit and PD, consistent with antagonistic pleiotropy in which alleles that increase liability to smoking initiation are associated with reduced PD risk. AKT3 functions within the PI3K-AKT-TrkB neurotrophic signaling pathway [55], while EXD2 regulates mitochondrial homeostasis and oxidative stress responses [56]. The latter is notable given that smoking-related neuroprotection may involve attenuation of mitochondrial oxidative stress via inhibition of MAO-B, which is suppressed by components of tobacco smoke and leaf extracts [16, 17]. Additionally, low-dose CO exposure has been proposed to protect against oxidative stress and neurodegeneration in preclinical PD models [20, 21]. The present findings converge with these previously proposed candidate protectants, providing genetic and proteomic context for mitochondrial stress-response pathways that have been hypothesized to underlie smoking-related neuroprotection. In contrast, RIT2 and MAPT showed concordant effects, aligning with their established roles in synaptic vesicle trafficking and cytoskeletal integrity [62, 86]. Together, the presence of both discordant and concordant proteins highlights a mixture of antagonistic and shared pathways that connect smoking liability and PD and demonstrates how multi-omic integration can refine mechanistic targets relevant to PD risk.

Finally, our conjFDR analyses identified multiple biologically tractable therapeutic targets with established interactions with FDA-approved medications. Neurotrophic potentiation strategies could enhance TrkB/PI3k-AKT signaling to promote dopaminergic neuron survival and resilience using NTRK2/TrkB signaling modulators such as larotrectinib and entrectinib. Microglial modulation via *P2RY12* could bias the immune response toward neuroprotection using drugs such as clopidogrel and ticagrelor. This immune mechanism is consistent with prior work showing smoking-gene interactions in PD involving *HLA-DRB1*, and with recent studies linking P2RY12 signaling to MHC class II-associated microglial activation states [87–90]. Other medications include aducanumab and newer generation amyloid immunotherapy targeting *APP*/amyloid biology, fampridine and amifampridine targeting *KCNH5*, lansoprazole targeting *MAPT* and tau regulation, and selinexor targeting *XPO1* nuclear exporter can also be used to test pathway-specific neuroprotection in PD without the systemic harms of tobacco exposure. Collectively, our findings highlight clinically actionable and mechanistically-grounded targets that bridge genetic risk and pharmacologic tractability.

This study has limitations. Residual confounding from unmeasured pleiotropy or subtle population structure cannot be excluded. The smoking phenotypes are complex and heterogeneously measured, and CigDay remains vulnerable to index-event biases and was less well-powered than SmkInit. Furthermore, our tissue-expression analyses relied on bulk tissue data, which lacks cell-type resolution. Moreover, our analyses only included individuals of European ancestry, which may limit generalizability to other populations. Despite these limitations, the study provides convergent evidence from multiple complementary genetic and proteomic approaches applied across large-scale, datasets. Collectively, these methods provide robust support for our finding of a protective effect of genetic liability to smoking initiation on PD risk

Future work should prioritize cross-ancestry fine-mapping to pinpoint causal variants, coupled with colocalization analyses using single-cell expression data from the midbrain to link these variants to specific cell types (e.g., dopaminergic neurons, microglia, astrocytes). Transcriptome- and proteome-wide MR could help to identify the specific genes and proteins that mediate these effects. Ultimately, functional validation is essential and will likely require complementary experimental systems. Human induced pluripotent stem cell-derived dopaminergic neuronal and microglial co-culture systems provide a platform to perturb cell-specific and intercellular candidates like *P2RY12* and *NTRK2* [91, 92]. In parallel, in vivo α-synuclein-based models such as transgenic and preformed fibril paradigms offer a framework to assess how modulating these pathways influences synuclein pathology, neuroinflammation, and neuronal survival [93, 94]. Integrating these insights across models will be a critical step in translating these genetic signals into tangible therapeutic strategies.

In summary, our integrated genetic and proteomic analysis provides a compelling model for the long-observed inverse association between smoking and PD. We identify variants that appear to confer modest protection against PD by modulating neurotrophic, synaptic, and microglial pathways active in PD-relevant brain circuits. By decoupling these protective biological pathways from the harmful effects of smoking, our findings offer a clear and rational basis for the development of novel, tobacco-free neuroprotective therapies for PD.

## Supporting information

Supplemental results

## Data Availability

All data produced in the present work are contained in the manuscript.

## Disclaimer

The contents of this publication are the sole responsibility of the authors and do not necessarily reflect the official policy or position of the Uniformed Services University of the Health Sciences, the Department of War, or Henry M. Jackson Foundation for the Advancement of Military Medicine, Inc.

## Conflict of Interest Disclosures

Dr. Kranzler is a member of advisory boards for Altimmune and Clearmind Medicine; a consultant to Sobrera Pharmaceuticals, Altimmune, Lilly; and Ribocure; and the recipient of research funding and medication supplies for an investigator-initiated study from Alkermes and company-initiated studies by Altimmune and Lilly.

## Funding

Dr. Aliza Wingo is supported by grant I01BX005686. Dr. Thomas Wingo is supported by R01 AG075827. Dr. Kranzler is supported by the Veterans Integrated Service Network’s Mental Illness Research, Education and Clinical Center; U.S. Department of Veterans Affairs grant I01 BX004820 and NIAAA grant R01 AA030056. Dr. Gray is supported by NIAAA grant R01 AA030041 and Department of War grant HU0001-22-2-0066.

## Notes

### Author Declarations

The source data are openly available

