## Supplemental results for "Genetic and Proteomic Investigation of the Smoking-Parkinson’s Disease Association"

**Shared Genomic Architecture between CigDay and PD**

Univariate MiXeR models produced finite AIC values, reflecting good model fit and adequate power for subsequent bivariate analyses. MiXeR analysis revealed a mild inverse genetic relationship between CigDay and PD (r_g_ = -0.092), with a polygenic overlap quantified by a Dice coefficient of 22.9%. Specifically, out of an estimated 1,774 potential causal variants linked to CigDay and 994 associated with PD, approximately 320 variants were shared. Notably, 34.8% of the shared causal variants displayed concordant effect directions (Table S2).

Conditional quantile-quantile (Q-Q) plots demonstrated cross-trait genetic enrichment between CigDay and PD, further supporting shared polygenic architecture. In these analyses, SNPs were stratified by their association with the primary trait (e.g., PD) at increasingly stringent p-value thresholds (*P* ≤ 0.1, 0.01, and 0.001). As shown in the conditional Q-Q plots, SNPs more strongly associated with PD exhibited progressively greater enrichment for association with CigDay, illustrated by an increasingly leftward shift from the null line. The same reciprocal enrichment pattern was observed when stratifying by CigDay significance and evaluating PD associations. These non-random enrichment patterns confirm a subset of shared genetic variants influencing both traits, reinforcing the modest but meaningful polygenic overlap detected by MiXeR (Figure S6).

**Shared Genetic Loci (cond/conjFDR)**

At conjFDR < 0.05, we identified 26 significant loci associated with both CigDay and PD. Half of the lead SNPs (50%) had concordant (consistent) effect directions for CigDay and PD, while the remainder were discordant (Table S4). The shared architecture span genes involved in dopaminergic neuron development, synaptic vesicle trafficking, lysosomal function, and calcium-dependent signaling (e.g., *SOX6*, *TMEM163*, *SCARB2*, *PPP3CC*, *EPHA5*, *PCDH17*), as well as epigenetic and RNA regulatory mechanisms (e.g., *SETD1A*, *PHF2*, *CXXC4*, *YTHDF3*). Additional genes relate to immune signaling and nicotine metabolism (*ITGAM*, *CNPY3*, *CYP2B6*), suggesting that smoking intensity and PD risk intersect across synaptic, lysosomal, transcriptional, and metabolic pathways (Table S4).

**FUMA Genotype-Tissue Expression analysis**

Analysis of Genotype-Tissue Expression (GTEx) data for genes linked to the 26 lead SNPs associated with both CigDay and PD did not show significant differential expression enrichment after Bonferroni correction at the level of 30 general tissues, and in the 54 specific-tissue panel (Figure S7).
